# Modeling effect of hypertension control on death, incidence of atrial fibrillation and economic impact to Medicare and hospitals

**DOI:** 10.64898/2026.07.15.26358198

**Authors:** Jeffrey Williams, Nicholas Mencer, Wen Yao Mak, Greta Dalle Luche, Sophie Dundovic

## Abstract

**Background:** Hypertension is a major modifiable risk factor for atrial fibrillation (AF), yet blood pressure (BP) control remains suboptimal in older U.S. adults.

**Objectives:** This study evaluated how improve systolic BP (SBP) control could affect incident AF, downstream AF ablation demand, Medicare savings, and hospital revenue.

**Methods:** A population-based modelling framework was developed to estimate mortality and incident AF hazards across SBP strata: <120, 120-139, 140-159, and ≥160 mm/Hg. AF incidence in the SBP <120 mmHg group was set at 2.2 per 1,000 person-year, with hazard ratios of 1.17, 1.42 and 1.64 applied to higher SBP strata. We assumed 25% of incident AF patients would undergo ablation, with a 7.2% complication rate. AF prevalence was projected to increase by 4.6% annually over 10 years. Medicare savings and hospital revenue foregone were estimated under varying procedure cost and contribution-margin assumptions.

**Results:** Higher SBP was associated with greater hazards of death and incident AF. Improved SBP control reduced projected AF incidence and ablation demand. Over 10 years, cumulative Medicare savings were projected at $8.7B-$10.9B across the full modelled population. However, reduced ablation volume translated into hospital revenue foregone, ranging from $75M to $377M in the first year, and approximately $1.03B-$5.2B cumulatively over 10 years.

**Conclusions:** Improved SBP control may reduce AF incidence, prevent avoidable invasive ablation procedures, relieve pressure on surgical waitlists, and generate substantial Medicare savings. However, these benefits may reduce hospital procedural revenue, highlighting a misalignment between prevention-oriented care and fee-for-service reimbursement incentives.

## Introduction

Hypertension (HTN) is one of the most prevalent and significant modifiable risk factors for atrial fibrillation (AF) in the general population^1^. In the United States, HTN affects approximately 77.1% of individuals aged >65 years and is present in 60-80% of patients with AF^2,3^. Among known risk factors, HTN accounts for the largest proportion of AF cases^4^ and remains the leading cause of death worldwide^5^. Pathophysiologically, HTN contributes to left ventricular hypertrophy and left atrial enlargement, creating an anatomical substrate conducive to AF development^6^.

Despite its importance, blood pressure control among the patients at risk of AF development remains suboptimal. Only 36.8% of adults aged >75 years achieve a target blood pressure of <140/90 mmHg, and fewer than 20% receive appropriately intensified antihypertensive therapy. A Mendelian randomization study involving 157,902 participants demonstrated that hypertension increases the risk of incident AF by approximately 50%, supporting a causal relationship between elevated blood pressure and AF^7^. Improving treatment intensification and adherence thus represents a major opportunity to reduce hypertension-related morbidity and mortality in the United States^8,9^.

This gap in HTN control reflects a deeper systematic failure of implementation of knowledge. In a recent Editor’s Page of JACC, Krumholz argued that severe uncontrolled HTN should be regarded as a modern “never-event”, as preventable and unacceptable as wrong-site surgery or fatal medication errors^10^. The persistence of untreated or undertreated HTN represents a systemic breakdown in detection, treatment, and accountability, with substantial downstream consequences including AF, stroke, heart failure, and premature death.

Most clinical studies have repeatedly demonstrated a linear association between systolic blood pressure (SBP) and AF incidence^4,11^. However, the impact of intensive BP lowering on AF risk remains uncertain due to limited published evidence^4^. To address this gap, we developed a population model to evaluate the potential effects of intensive SBP reduction on AF incidence and overall mortality. We also estimated the extent to which intensive SBP control could reduce the demand for AF ablation and evaluated the associated economic implications for Medicare and hospital systems.

## Methods

### Model-based hazard estimations of mortality and incident atrial fibrillation associated with treatment of hypertension

A model-based framework was developed in MATLAB (MathWorks Inc, Natick, MA) to estimate the hazards of all-cause mortality and incident AF under varying levels of blood pressure control. Two models were developed separately to estimate the hazard of AF and mortality. The models assumed that cumulative survival times for these outcomes followed an exponential survival distribution, in which the hazards (event of AF development and mortality) rate remained constant over time^12^. Parameterization of the hazard functions was informed by data from major randomized and observational hypertension trials, including the HYVET Study Group^13^, the SPRINT Research Group^14^, the Korean National Health Insurance Service cohort^15^, and the ONTARGET/TRANSCEND studies^11^. These sources collectively provided population-level estimates linking SBP to risks of mortality and new-onset AF across diverse cardiovascular risk strata.

Specifically, model inputs and parameter values for mortality and AF hazards were derived from published event rates and treatment effects reported in these studies. These parameters were used to estimate the hazard of death as a function of SBP (as shown in Table 1), and the corresponding parameters for estimating incident AF based on SBP and overall cardiovascular (CV) risk profiles were tabulated in Table 2.

**Table 1.**
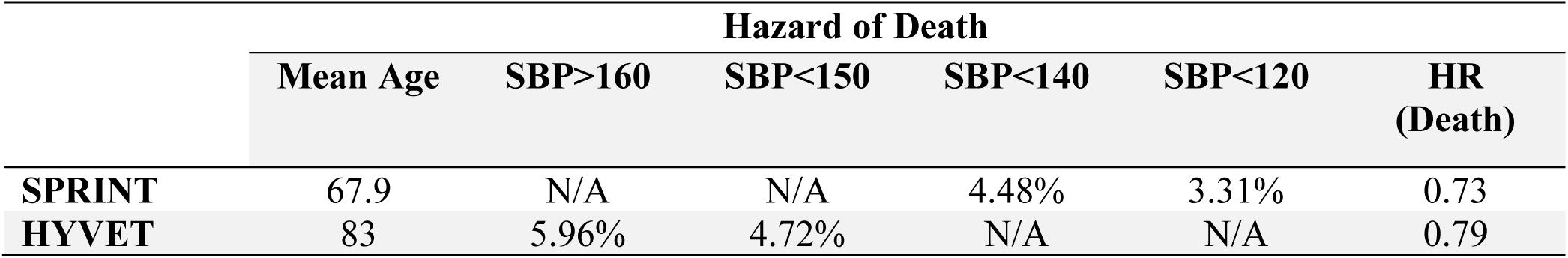
Effect higher SBP has upon hazard of death.

**Table 2.**
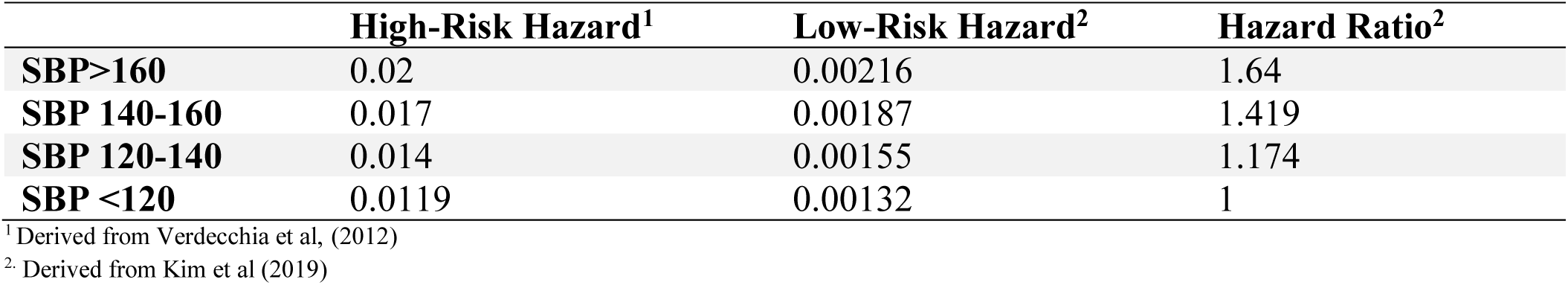
Cumulative Incidence Hazards of Atrial Fibrillation and SBP.

### Modeling impact of SBP control on future AF ablation demand

A subsequent population-based economic projection model was developed to estimate the 10-year impact of improved SBP control on the incidence of AF and the associated healthcare costs in the hypertensive U.S. population aged more than 65 years. The model aimed to incorporate published epidemiologic data and national demographic statistics to reflect real-world population characteristics.

Hypertensive individuals were stratified by SBP into group categories: *normal* (<120 mmHg), *stage 1* (120-139 mmHg), *stage 2* (140-159 mmHg), and *most vulnerable* (≥160 mmHg). The baseline AF incidence among the normal SBP group was assumed to be 2.2. cases per 1,000 person-years, with corresponding hazard ratios (HRs) of 1.17, 1.42, and 1.64 for progressively higher SBP strata based on the pooled cohort analyses linking SBP to new-onset AF (see Table 2). These HRs were applied to estimate incident AF cases under both current and optimized BP control conditions. We assumed approximately 25% of newly diagnosed AF patients^16^ would undergo ablation based on existing literature, consistent with procedural trends among Medicare beneficiaries. The procedure-related complication rate was set at 7.2% encompassing both periprocedural and readmission-related events^17^. The prevalence of AF was projected to increase by 4.6% annually over the next 10 years, in alignment with the national epidemiologic projections in the U.S.^18,19^

### Modeling economic impacts of reduced AF ablation demand on Medicare and hospitals

Economic parameters were derived from Medicare reimbursement data and hospital cost reports to estimate financial outcomes associated with AF management under different BP control scenarios. For fiscal year (FY) 2025, the Medicare Severity Diagnosis-Related Group (MS-DRG) reimbursement rates for catheter ablation procedures were $27,906 for MS-DRG 273 (“percutaneous and other intracardiac procedures with major complications or comorbidities [MCC]”) and $22,211 for MS-DRG 274 (“without MCC”). The model used a weighted average reimbursement of approximately $26,000 per ablation to reflect the national payer mix.

An incremental hospitalization cost of $7,812 was applied to cases involving procedural complications, based on published Medicare costs analyses^20^. Hospital contribution margins, defined as the percentage of revenue remaining after variable costs are subtracted from total revenue, were modelled at 10%, 30%, and 50% that represented *low, moderate*, and *high profitability* tiers respectively. Two policy scenarios were evaluated based on the framework. The “status quo” scenario represented the current population distribution of SBP, whereas the “improved control” scenario simulated a population-level shift toward lower SBP categories that aligned with successful antihypertensive therapy. For each scenario, annual and cumulative differences in AF incidence, ablation volume, and total cost burden were projected over a 10-year horizon. Medicare savings were estimated under three pricing tiers corresponding to the contribution margin levels, while hospital revenue foregone was calculated based on reductions in ablation volume under improved BP control. Sensitivity analyses were performed to evaluate the robustness of results to variations in BP reduction magnitude, ablation uptake rates, and hospital margin assumptions.

All model parameter assumptions and input values are summarized in Table 3 to ensure transparency and reproducibility.

**Table 3:** Model parameters and assumptions.

|  |  |
| --- | --- |
| Numbers in millions, n (%) | Subgroup population |

| Hypertension classification | SBP <120<br>mmHg | SBP 120-140<br>mmHg | SBP 141-160<br>mmHg | SBP >160<br>mmHg |
| --- | --- | --- | --- | --- |
| Estimated HTN population | 8.932 | 8.932 | 13.398 | 13.398 |
| Estimated incident AF<br>from HTN | 19650.4 | 22990.9 | 41855.4 | 48339.9 |
| Estimated AF patients who<br>required ablation | 4912.6 | 5747.742 | 10463.838 | 12084.996 |

## Results

### Model-based hazard estimations of mortality and incident AF associated with treatment of HTN

The cumulative hazard of death increased progressively with higher SBP levels and cohort age. Figure 1A shows that an SBP increase of 10-20mmHg carries a relative risk of 1.55-1.78, potentially heightened by patient risk, as a higher SBP led to a greater relative risk in those with a higher baseline CV risk. Among the older adults (mean age of 83 years, HYVET Study Group), patients within the *most vulnerable* SBP group (>160 mmHg) had the highest cumulative hazard of approximately 0.25 in the fifth year. In the comparatively younger cohort (mean age 67.9 years, SPRINT Research Group), the cumulative hazard was approximately 0.18 for SBP < 140 mmHg and 0.14 for SBP < 120 mmHg.

**Figure 1.**
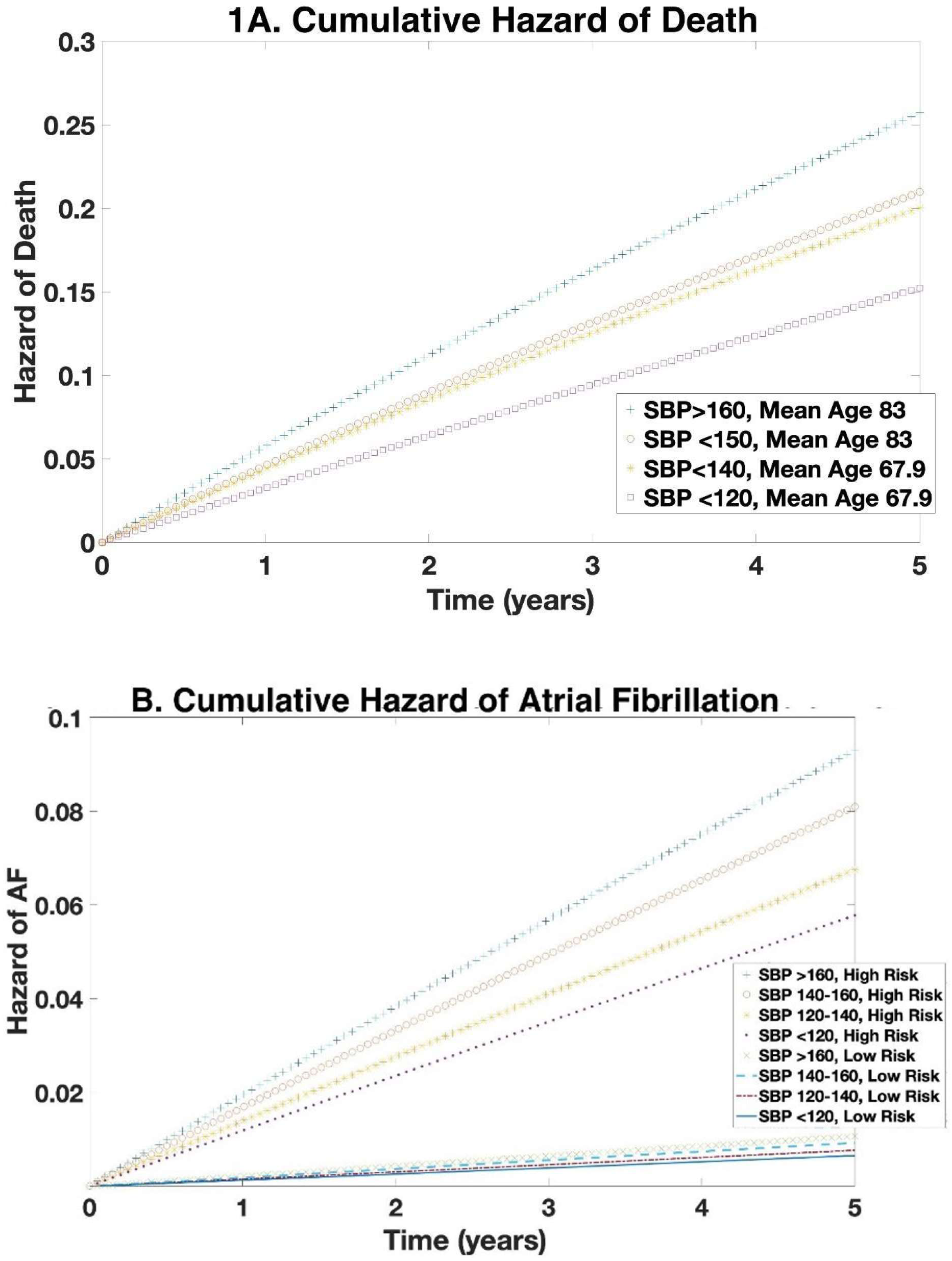
Figure A depicts the effect higher SBP has upon hazard of death. Figure B depicts the effect of SBP on the cumulative hazard of incident AF.

Figure 1B shows that regardless of level of cardiovascular risk, from a baseline SBP<120, a SBP 120-140, 140-160, and >160 resulted in a relative risk of incident AF of 1.174, 1.419, and 1.64, respectively. Under current conditions, approximately 113,000 new AF cases occur annually among hypertensive and pre-hypertensive (>120 mmHg) adults aged ≥65 years. Using the established effect of BP control on AF incidence, we estimated the reduction in ablation procedures achievable with effective hypertension management in US patients. Treating only the most vulnerable group to target BP was projected to prevent roughly 12,000 AF ablations during the first year, saving Medicare $328M in avoided procedure and complication costs. Expanding treatment to those stage-2 hypertensive patients further reduced AF incidence, resulting in $613M in Medicare savings. Comprehensive BP control across all hypertensive categories (SBP >120 mmHg) prevented the greatest number of AF cases, corresponding to $770M in Medicare savings.

### Modeling impact of SBP control on future AF ablation demand and its economic impacts for Medicare and hospitals

Over ten years, projected cumulative Medicare savings ranged $8.7B-$10.9B (all patients, Figure 2. Note the figure demonstrates yearly saving). Hospital revenue foregone ranged $75M (10% margin)-$377M (50% margin) for the first year only (see Figure 3) leading to an estimated cumulative revenue loss of $1.03B (10% margin) to $5.2B (50% margin) over ten years (calculated using the medium cumulative savings of $10.4B).

**Figure 2.**
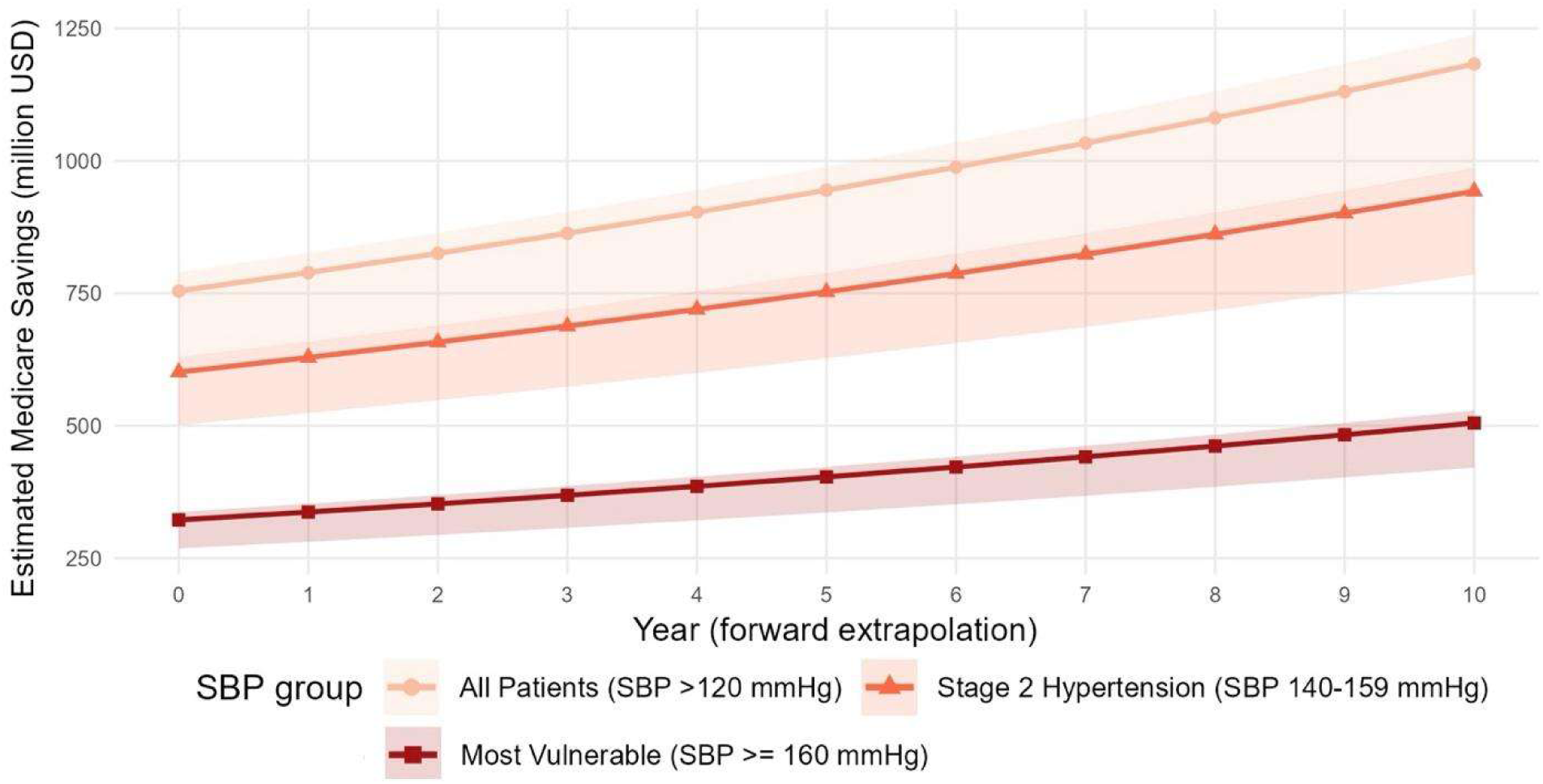
Estimated Medicare savings based upon strata of blood pressure control.

**Figure 3.**
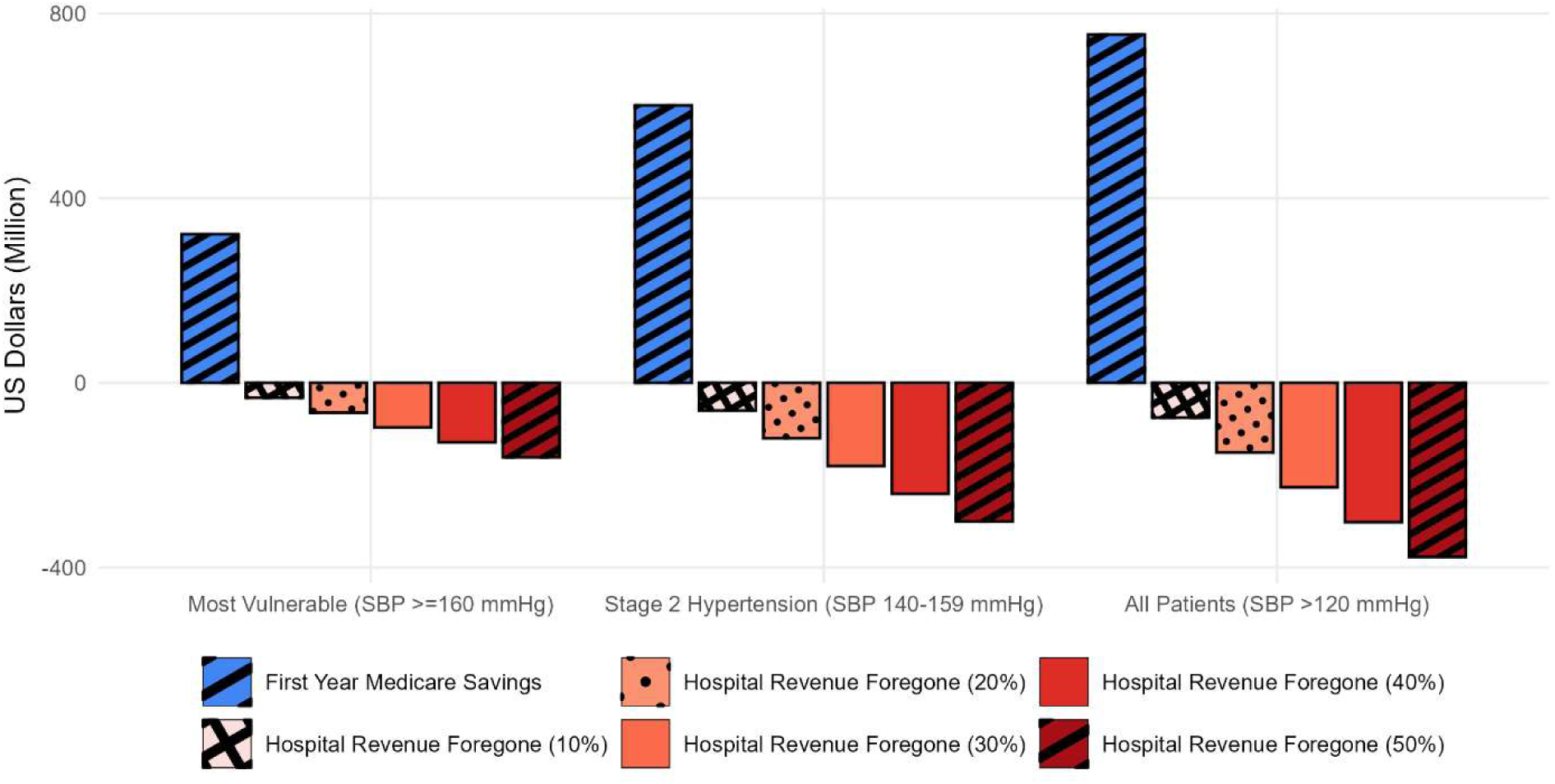
Medicare savings versus hospital revenue based upon degree of blood pressure control.

## Discussion

The prevalence of both HTN and AF are growing, and studies supported the linear relationship between SBP and AF^4,11^. Our model suggests that intensive BP control confers substantial clinical and economic benefits, including reductions in cumulative mortality, new-onset AF, and a marked decrease in ablation procedures among U.S. Medicare beneficiaries. Over a ten-year horizon, projected Medicare savings ranged from $985 million to $1.23 billion, driven primarily by lower AF incidence and procedure volume.

Beyond the economic effects, improved SBP control may also have important benefits for patients and healthcare delivery. Guideline-driven intensification of HTN management^21^ could reduce the number of patients progressing to AF that require ablation, thereby avoiding invasive procedures and related complications that necessitate further exhaustion of healthcare resources. At the systemic level, efforts to lower demands for ablation could help to relieve pressure on surgical waitlists and preserve the surgical capacity for patients with more severe arrhythmia or fibrillation. In this context, strengthening SBP control is not only a preventive strategy, but it is also a possible mechanism to improve healthcare resource allocations and cardiovascular care.

In addition, these same benefits translated into foregone hospital profits ranging from $75 million (assuming a 10% margin) to $377 million (assuming a 50% contribution margin), underscoring a structural misalignment between payer and provider incentives within the current fee-for-service reimbursement framework. This paradox highlights a broader challenge within the U.S. healthcare system, in particular, the improvements in preventive cardiovascular care can diminish procedural revenue even as they enhance population health outcomes. Such dynamics may partly explain why national healthcare expenditures have reached unprecedented levels while cardiovascular morbidity and mortality continue to worsen^22^. The U.S. population exhibits an exceptionally high burden of poorly controlled cardiometabolic risk factors, with 90-95% of adults having at least one clinically elevated risk factor (most commonly HTN)^23^. Unfortunately, heart failure mortality appears to be on the rise, fueled by concurrent epidemics of obesity, diabetes, and uncontrolled BP^24^. Among traditional risk factors, SBP remains the single most modifiable determinants of cardiovascular disease risks^25^. Evidence from randomized trials demonstrates that effective HTN treatment reduces the risk of stroke by 15-25%, myocardial infarction by 15-25% and heart failure by up to 64%^26–28^.

Our findings further contextualize AF as both a clinical and economic inflection point in cardiovascular care. AF is among the leading contributors to hospital revenue and Medicare spending, particularly through rhythm-control interventions such as catheter ablation. Randomized trials have shown that rhythm control with antiarrhythmic drugs (such as EAST-AFNET4^29^, ATHENA post-hoc analysis^30^) and management of underlying disease drivers (see Table 3) improve hard cardiovascular outcomes, whereas AF ablation has yet to demonstrate consistent benefits on mortality or major adverse cardiovascular events despite effectiveness at reducing AF burden (CABANA^31^, STOP-AF^32^, RAFT-AF^33^). The present model illustrates the paradox wherein suboptimal HTN control can indirectly sustain hospital revenue streams, creating an economic disincentive for aggressive BP management.

From a pathological perspective, HTN is the predominant upstream cause of AF in the Medicare population, yet a growing subset of AF cases occurs in the absence of overt HTN, chronic heart failure, or valvular disease that cause hemodynamic stresses on the left atrium^6^. These patients often exhibit atrial myopathy, with AF being the most clinically evident manifestation. Underlying systemic inflammatory disease leads to epicardial adipose tissue expansion and inflammation causing an enlarged fibrotic and noncompliant left atrium (LA)^34^. It becomes evident that AF is a marker for a broad range of poorly controlled underlying systemic inflammatory and metabolic diseases beyond HTN and ablation is a focal solution to a systemic problem. Our model focused exclusively on HTN, but with the potential to expand into various other modifiable systemic disorders becomes the target of AF management (rather than relying on ablation, see Table 4).

**Table 4.** Underlying systemic disease processes that affect atrial fibrillation.

| **Modifiable Disease Process** | **Pathologic Mechanisms** | **Level of Undertreatment/Underdiagnosis** | **Effect on AF** | **Treatment Effects on CV Outcomes** |
| --- | --- | --- | --- | --- |
| Hypertension | Increased LV afterload and peripheral vascular resistance leading to LV structural remodeling and diastolic dysfunction<sup>35</sup>. | Less than 20% of adults with appropriate intensification of therapy<sup>36</sup>. | Doubles the risk for AF and accounts for more AF than any other risk factor<sup>37</sup>. | BP lowering associated with 31% RRR stroke and 22% reduction in MACE<sup>38</sup>. |
| Diabetes | Accumulation of advanced glycation end products, oxidative stress, inflammation, decay of intracellular calcium<sup>39,40</sup>, and progression of atherosclerotic vascular disease<sup>41</sup>. | 16% diabetics undiagnosed; 20% of diabetics poorly controlled<sup>42,43</sup>. | Associated with 1.6-fold increased risk of AF<sup>19</sup>. | Empagliflozin gives 37% RRR in death from CV causes<sup>44</sup>. |
| Obesity | Sustained obesity leads to inflammation, hypertension, DM, HTN, OSA, decay of intracellular calcium<sup>39,40</sup> metabolic syndrome, and coronary heart disease<sup>19</sup>. Hemodynamic changes include increased plasma volume, LV remodeling, LA volume, RV dysfunction, and epicardial fat<sup>45</sup> | Over half of all adults are overweight or obese<sup>46</sup>. | Obesity increases the risk of developing new-onset AF by 50% and is the strongest predictor for developing AF following hypertension<sup>46</sup>. | Weight loss leads to 48% reduction in risk of death<sup>12</sup>. Weight loss has demonstrated success in both primary (29% lower incidence) and secondary prevention (6-fold greater freedom) of AF<sup>47,48</sup>. |
| Atherosclerotic | Two-thirds of heart failure cases are attributed to | In patients with established ASCVD, only | Atherosclerosis is associated with new onset | Statins can reduce the risk of ASCVD events by |
| Cardiovascular Disease | underlying coronary artery disease <sup>49</sup>. | 22.5% of patients on a high-intensity statin, 27.6% were on a low- or moderate-intensity statin, and 49.9% were not on any statin <sup>50</sup>. | AF in patients without established CAD<sup>51</sup>. MI is a strong predictor of AF especially in men <sup>52</sup>. | approximately 30% with an additional 15% reduction with high-intensity compared to low- or moderate-intensity regimens <sup>50</sup>. |
| Chronic Kidney Disease | Insulin resistance, sodium/fluid retention, increased RAAS/oxidative stress/endothelial dysfunction/vascular calcification<sup>53</sup> | 14% of US adults have CKD (eGFR<60mL/min or UACR>30). 60% of people with CKD are missed because UACR not checked in patients with normal eGFR but at risk (due to HTN or DM2) <sup>54</sup>. | CKD is one of the strongest risk factors for CVD manifest as CAD <sup>53</sup>. 90% of patients with CKD have HTN. CKD is also associated with increased risk of new-onset AF<sup>55</sup> | Statins reduce CV events in CKD. 10% reduction in weight slows progression of CKD/albuminuria. ARB/SGLT2/MRA/GLP1 agonists improve CV outcomes and slow CKD progression <sup>53</sup>. |
| Heart Failure | Hypertension, obesity, diabetes, and atherosclerotic cardiovascular disease impart the highest relative risks and population attributable risk for development of HF <sup>56</sup>. | The lifetime risk of heart failure has been estimated at 20-46% <sup>57</sup>. Shockingly, less than 1% of patients with heart failure are receiving all life-prolonging treatments at trial-proven doses <sup>58</sup>. | AF development is 4-6x higher in patients with heart failure <sup>52</sup>. | Beta blockers, MRA, SGLT-2 inhibition and Angiotensin-Neprilysin Inhibition reduce mortality and CHF readmission <sup>59</sup>. |
| Inflammation | Chronic, silent, low-grade inflammation, together with key mediators like cytokines, chemokines, and acute-phase reactants, plays a pivotal role in HTN, atherosclerosis and AF <sup>60-62</sup>. | The prevalence of a moderate to higher risk hsCRP levels $\geq 2$ mg/L in adults in the primary prevention setting may vary from approximately 30% to 35% in Europe to 50% in the United States <sup>61</sup>. | The link between high hsCRP and future CVD is as strong as the link between HTN and CVD; this connection appears to be more significant than seen with traditional lipids <sup>63</sup>. | Evolving anti-inflammatory approaches to secondary prevention: colchicine 20-25% reduction in CV risk, canakinumab 15-17% reduction in vascular events, lifestyle modification (smoking cessation, weight loss, exercise), and low-level vagus nerve stimulation<br>has resulted in 85%<br>reduction in burden of AF<br>61,64–66 |

From a policy and health system standpoint, these findings underscore the urgent need to realign financial incentives toward prevention. Fee-for-service payment structures rewards procedural volume rather than disease risk reduction, thereby discouraging investments in long-term cardiometabolic control. Transitioning toward value-based reimbursement that is anchored on risk reduction and quality-adjusted life-years gained could harmonize clinical and economic objectives. Preventive cardiology that focuses on optimized management of hypertension and related metabolic disorders represent one of the most cost-effective strategies to reduce AF burden and mortality in the aging U.S. population.

## Limitations

Our work focused exclusively on the effect of HTN control, though it is likely that both the clinical and economic benefits would be amplified if additional systemic diseases were incorporated. Important comorbidities such as diabetes, obesity, heart failure and inflammatory disorders should be explored further. The model also assumed that 25% of patients with atrial fibrillation underwent ablation, based on data from a large, multicenter, prospective cohort reporting a 21% prior ablation rate. While ablation rates vary across regions and healthcare systems, we do not expect such variability to alter the fundamental relationship between HTN control, Medicare savings, and hospital revenue loss. The analysis did not account for potential cost savings related to reductions in AF-related hospitalizations resulting from lower AF incidents. Furthermore, the model assumed a certain level of adherence to antihypertensive therapy, which may not reflect real-world patterns. Notably, approximately 24-28% of U.S. adults aged >65 years with hypertension are estimated to be nonadherent to their prescribed BP regimes^67^. This limitation suggests that the projected benefits of improved BP control could be conservative if adherence were optimized.

## Conclusion

Our modeling indicates that improved management of HTN improves survival and reduces incident AF. This improvement in patient health leads to fewer ablations and substantial savings for Medicare over ten years. This scenario also highlights a problem with the current U.S. healthcare reimbursement system. The resulting reduction in hospital profits from improved patient care may disincentivize prevention. This dilemma may help explain the rising rate of heart failure deaths despite historically high healthcare spending, particularly as traditional risk factors like hypertension, obesity, and diabetes are poorly managed across the country. Optimal management of HTN is seen as having the greatest potential for preventing cardiovascular disease.

## Data Availability

All data used in this study were extracted from publicly available, previously published sources.

## Credit Author Statement

**Jeffrey William:** Conceptualization, Methodology, Writing - original draft, Writing - review & editing **Nicholas Mencer:** Data curation, Methodology, Software, Formal analysis. **Wen Yao Mak:** Formal analysis, Visualization, Writing - original draft, Writing - review & editing. **Greta Dalle Luche:** Investigation, Writing - review & editing. **Sophie Dundovic:** Supervision, Funding acquisition, Resources.

